# Estimating body fat distribution – a driver of cardiometabolic health – from silhouette images

**DOI:** 10.1101/2022.01.14.22269328

**Authors:** Marcus D. R. Klarqvist, Saaket Agrawal, Nathaniel Diamant, Patrick T. Ellinor, Anthony Philippakis, Kenney Ng, Puneet Batra, Amit V. Khera

## Abstract

**Background:** Inter-individual variation in fat distribution is increasingly recognized as clinically important but is not routinely assessed in clinical practice because quantification requires medical imaging.

**Objectives:** We hypothesized that a deep learning model trained on an individual’s body shape outline – or “silhouette” – would enable accurate estimation of specific fat depots, including visceral (VAT), abdominal subcutaneous (ASAT), and gluteofemoral (GFAT) adipose tissue volumes, and VAT/ASAT ratio. We additionally set out to study whether silhouette-estimated VAT/ASAT ratio may stratify risk of cardiometabolic diseases independent of body mass index (BMI) and waist circumference.

**Methods:** Two-dimensional coronal and sagittal silhouettes were constructed from whole-body magnetic resonance images in 40,032 participants of the UK Biobank and used to train a convolutional neural network to predict VAT, ASAT, and GFAT volumes, and VAT/ASAT ratio. Logistic and Cox regressions were used to determine the independent association of silhouette-predicted VAT/ASAT ratio with type 2 diabetes and coronary artery disease.

**Results:** Mean age of the study participants was 65 years and 51% were female. A deep learning model trained on silhouettes enabled accurate estimation of VAT, ASAT, and GFAT volumes (R^2^: 0.88, 0.93, and 0.93, respectively), outperforming a comparator model combining anthropometric and bioimpedance measures (ΔR^2^ = 0.05-0.13). Next, we studied VAT/ASAT ratio, a nearly BMI- and waist circumference-independent marker of unhealthy fat distribution. While the comparator model poorly predicted VAT/ASAT ratio (R^2^: 0.17-0.26), a silhouette-based model enabled significant improvement (R^2^: 0.50-0.55). Silhouette-predicted VAT/ASAT ratio was associated with increased prevalence of type 2 diabetes and coronary artery disease.

**Conclusions:** Body silhouette images can estimate important measures of fat distribution, laying the scientific foundation for population-based assessment.

## INTRODUCTION

Body-mass index (BMI) is a routinely measured proxy for overall fat burden. Increased BMI – used to define obesity in clinical practice – is a leading risk factor for cardiovascular disease, type 2 diabetes, and all-cause mortality.^1–4^ While BMI is a useful guide for disease risk at a population level, individuals with the same BMI can have markedly different fat distributions and cardiometabolic risk profiles.^5–8^ Prior work utilizing medical imaging such as magnetic resonance imaging (MRI), computed tomography (CT), and dual-energy X-ray absorptiometry (DEXA) has identified certain fat depots as key drivers of “within BMI-group variation” in cardiometabolic risk.^9,10^ At any given BMI, increased visceral adipose tissue (VAT) has been associated with cardiometabolic risk while abdominal subcutaneous adipose tissue (ASAT) may have a net neutral effect, and gluteofemoral adipose tissue (GFAT) appears to be a protective ‘metabolic sink’ for excess adipose tissue.^11–14^

These findings suggest potential value in quantifying fat depot volumes – either for identifying high-risk individuals based on metabolically unfavorable characteristics or for tracking response to a given weight reduction therapy. Based on the increased risk conferred by visceral fat, recent professional society guidelines suggest inclusion of waist circumference as a ‘vital sign’ within clinical practice.^15^ This is supported by the observation that waist circumference is correlated with VAT volume as well as a strong association of waist circumference with cardiovascular and all-cause mortality.^13,16–18^ However, for any given individual, waist circumference may be driven by fat surrounding internal organs (VAT) or fat accumulation just under the skin (ASAT), with potentially important differences in corresponding risk.

Hence, a large gap exists between anthropometric measures such as BMI and waist circumference – easily quantified in clinical practice, but providing limited resolution – and medical imaging, which allows for more precise characterization of fat distribution, but has not been practical to deploy at scale. Images of an individual’s silhouette – if adequately predictive – could close this implementation gap. Prior seminal studies have suggested that estimation of fat distribution using various proxies for medical imaging may be feasible, but have several limitations.^19–28^ First, most studies in this area have focused on predicting overall fat and fat-free mass rather than specific fat depots, which is likely to be significantly more diffcult.^19–23,28^ Second, no prior study has aimed to predict ratios between fat depots, which are poorly captured by BMI and waist circumference and hence may add the most clinical value.^24–27^ Third, prior studies in this area have been limited by sample sizes of up to several hundred healthy participants, limiting the ability to perform robust cross-validation. Fourth, no prior study has demonstrated that fat distribution predicted by an individual’s outline stratifies risk of cardiometabolic disease independent of BMI and waist circumference.

In this study, we derived front- and side-facing silhouette images for 40,032 participants of the UK Biobank from raw MRI imaging data. Deep learning models trained on these images, using previously calculated whole-body MRI-estimated volumes as truth labels, demonstrated highly accurate estimation of VAT, ASAT, and GFAT volumes, significantly outperforming prediction achievable using anthropometric variables.^14^ Beyond measures such as waist circumference, we note that the VAT/ASAT ratio quantified using silhouette images is a strong predictor of both type 2 diabetes and coronary artery disease.

## METHODS

### Study population

All analyses were conducted in the UK Biobank, a prospective cohort study that recruited over 500,000 individuals aged 40-69 in the UK from 2006 to 2010. In this study, we analyzed 40,032 participants of the imaging substudy with fat depot volumes previously quantified using whole-body MRI.^14,29^ This analysis of data from the UK Biobank was approved by the Mass General Brigham Institutional Review Board and was performed under UK Biobank application #7089.

### Preparing silhouettes from whole-body magnetic resonance images

Whole-body MRI data was preprocessed as previously described.^14^ In short, whole-body MRIs were acquired in 6 separate series with varying resolutions which require preprocessing before merging into 3D volumes. We resampled each series to the highest available resolution (voxel = 2.232 × 2.232 × 3.0 mm^3^), de-duplicated overlapping regions and merged the six series into 3D volumes. The fat-phase acquisition was used to segment a 3D volume for each individual, as described in the **Supplementary Methods**.

In order to generate a “silhouette” encoding information only about the outline of an individual, pixel intensities were set to one if they were on the surface of the body and to zero otherwise in either the coronal or sagittal orientation. For example, a given pixel on a coronal two-dimensional projection represents the presence or absence of a segmented pixel in the anterior-posterior direction perpendicular to the coronal plane. Classifying pixels as belonging to either the body or the background, in a procedure known as segmentation, was performed on the two-dimensional axial images. The input to the deep learning models described below was two such silhouette images concatenated side-by-side – one coronal and one sagittal – and resized to 237 × 256 pixels.

### Deep learning to predict fat depot volumes using silhouettes

For predicting the target fat depot volumes, we employed the DenseNet-121 architecture as the base model.^30^ Additional information regarding deep learning architecture, parameters, and training procedure can be found in the **Supplementary Methods**. In brief, we constructed a hierarchical multi-task model with the coronal and sagittal silhouettes as input that jointly predicted visceral (VAT), abdominal subcutaneous (ASAT), and gluteofemoral (GFAT) adipose tissue volumes, and VAT/ASAT ratio **(Supplementary Methods**).

To avoid reporting overfit results and to ensure that all participants received an unbiased prediction, we employed a nested cross-validation approach. In this approach, the cohort is first split into five non-overlapping partitions and five models can then be trained using data from three partitions, then testing and validation is performed using the remaining two partitions (**Supplementary Figure S1**). For each model, predictions from the validation partition are unbiased and are collected to acquire predictions for all participants. We found that performing cross-validation within the partitions improved performance. The final prediction for each fold was reported as the mean-ensemble of the cross-validation models.

### Linear anthropometric models to benchmark performance

Sex-specific anthropometric models were generated by predicting each MRI-derived fat measurement using one of, or a combination of, age, weight, height, body mass index (BMI), waist circumference, hip circumference, waist-to-hip ratio (WHR), and five bioelectric impedance measurements commonly used for measuring body fat. We utilized the aforementioned nested-cross validation approach to generate predictions from these models. R^2^ and mean absolute error (MAE) are reported to compare performance of models. 95% confidence intervals for R^2^ were generated by bootstrapping with 1,000 resamples.

### Association with Cardiometabolic Diseases

The primary outcomes were prevalent and incident type 2 diabetes and coronary artery disease.^14^ Type 2 diabetes was defined on the basis of ICD-10 codes, self-report during a verbal interview with a trained nurse, use of diabetes medication, or a hemoglobin A1C greater than 6.5% before the date of imaging (**Supplementary Table S8A**). Coronary artery disease was defined as myocardial infarction, angina, revascularization (percutaneous coronary intervention and/or coronary artery bypass grafting), or death from CAD as determined on the basis of ICD-10 codes, ICD-9 codes, OPCS-4 surgical codes, nurse interview, and national death registries (**Supplementary Table S8B**).

Sex-stratified logistic regression models adjusted for age at time of imaging, imaging center, BMI, and waist circumference were used to test associations of silhouette-predicted VAT/ASAT ratio with prevalent disease. Cox proportional-hazard models with the same covariates were used to test associations of silhouette-predicted VAT/ASAT ratio with incident events. Finally, we used sex-stratified logistic regression models adjusted for the same covariates to determine the gradient in probability of prevalent disease across quintiles of silhouette-predicted VAT/ASAT ratio in BMI- and waist circumference-bins.

## RESULTS

### Silhouettes allow for accurate estimation of VAT, ASAT, and GFAT volumes

40,032 participants of the UK Biobank imaging substudy with VAT, ASAT, and GFAT volumes previously quantified on the basis of MRI were included.^14,29,31–33^ Mean age was 65 years, 20,597 (51%) were female, and 97% were white (**Table 1**). Coronal and sagittal silhouettes were generated for each participant by (1) segmenting the body outline in axial MRI acquisitions, (2) computing a surface map of the resulting segmentation volume, (3) projecting this 3-dimensional surface map into 2-dimensional images in the coronal (front-to-back) and sagittal (side-by-side) directions, and (4) converting pixel intensities into binary values, either zeros or ones (**Figure 1A**). These silhouettes were used as inputs to train a convolutional neural network (CNN) model to predict VAT, ASAT, and GFAT volumes using MRI-derived measurements as truth labels.^14^

**Table 1.** Characteristics of the study population.

|  | **Males**<br>**(N = 19,435)** | **Females**<br>**(N = 20,597)** |
| --- | --- | --- |
| Age at time of MRI (years) | 65.2 ± 7.7 | 63.8 ± 7.5 |
| **Race** |  |  |
| White | 18,773 (96.6) | 19,936 (96.8) |
| Black | 137 (0.7) | 192 (0.9) |
| East Asian | 112 (0.6) | 137 (0.7) |
| South Asian | 238 (1.2) | 133 (0.6) |
| Other | 175 (0.9) | 199 (1.0) |
| **Blood pressure** |  |  |
| Systolic blood pressure (mmHg) | 142.0 ± 17.4 | 135.8 ± 19.2 |
| Diastolic blood pressure (mmHg) | 80.6 ± 9.9 | 76.7 ± 10.0 |
| **Anthropometrics** |  |  |
| Weight (kg) | 83.8 ± 13.2 | 68.9 ± 12.7 |
| Height (cm) | 176.3 ± 6.6 | 162.8 ± 6.35 |
| Body-mass index (kg/m<sup>2</sup>) | 27.1 ± 3.8 | 26.1 ± 4.6 |
| Waist circumference (cm) | 94.6 ± 10.5 | 82.8 ± 11.6 |
| Hip circumference (cm) | 100.9 ± 7.3 | 100.9 ± 9.6 |
| Waist-to-hip ratio | 0.94 ± 0.06 | 0.82 ± 0.07 |
| **Fat Depot Volumes (quantified by MRI)** |  |  |
| Visceral (L) | 5.0 ± 2.3 | 2.6 ± 1.5 |
| Abdominal subcutaneous (L) | 5.9 ± 2.5 | 7.9 ± 3.3 |
| Gluteofemoral (L) | 9.3 ± 2.6 | 11.3 ± 3.2 |
| **Cardiometabolic diseases (at time of imaging)** |  |  |
| Type 2 diabetes | 1,264 (6.5) | 637 (3.1) |
| Coronary artery disease | 1,542 (7.9) | 414 (2.0) |

Values are reported as means ± SD or as count (percentage). Race was adjudicated on the basis of self-reported ethnic background at time of enrollment in the UK Biobank.

**Figure 1.**
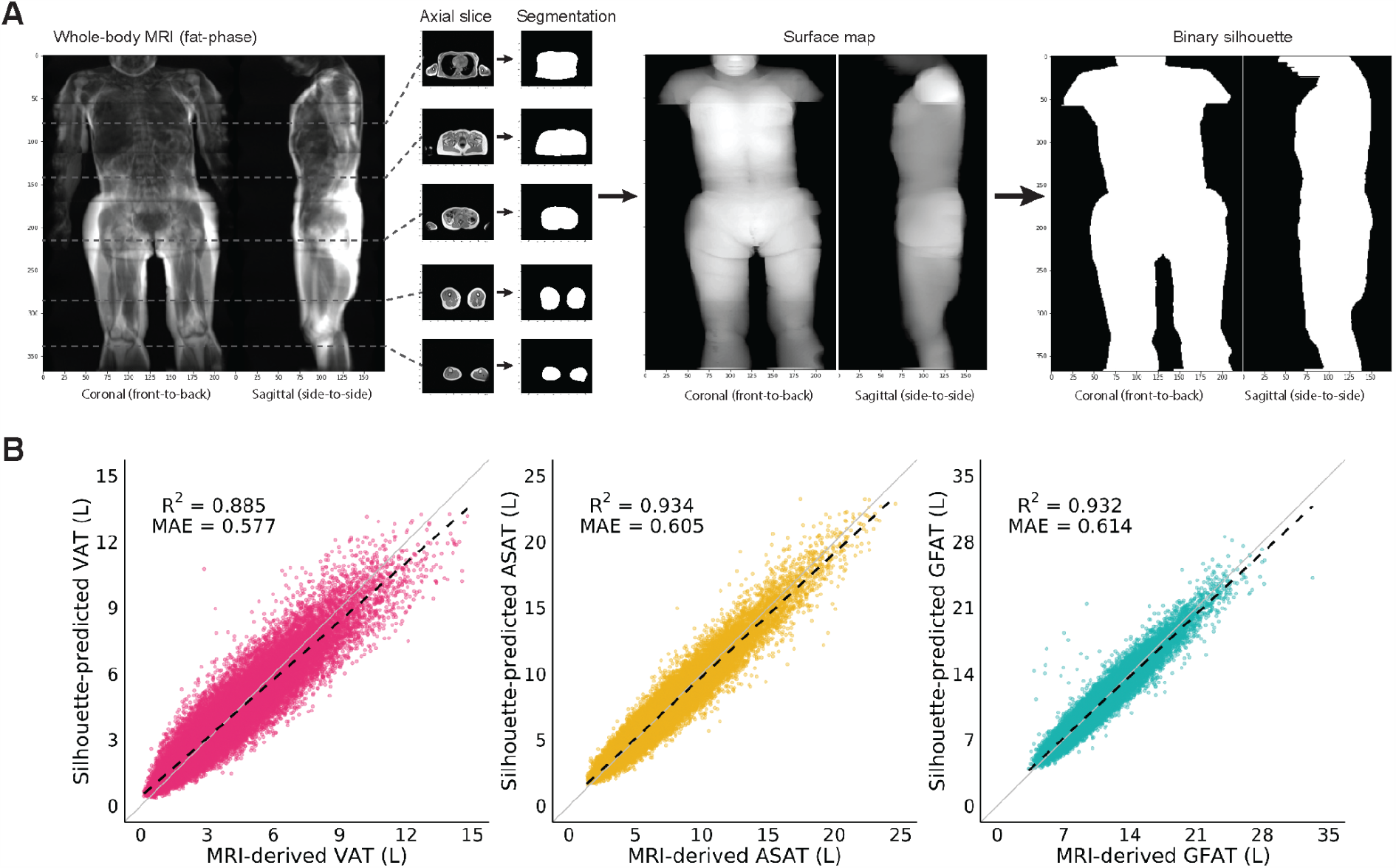
Silhouettes enable estimation of fat depot volumes. **A)** Silhouettes were created from body MRIs by segmenting the outline of axial acquisitions, projecting the resulting volume onto a 2-dimensional surface map, and binarizing pixels. **B)** Silhouette-predicted VAT, ASAT, and GFAT plotted against MRI-derived measurements.^14^ Solid black lines denote the linear fits, while the solid gray lines correspond to the identity line. Sex-stratified performance metrics and Bland-Altman plots are available in **Supplementary Table S1** and **Supplementary Figure S2-5**.

The CNN trained on silhouettes achieved high performance for predicting VAT (R^2^ = 0.885; 95%CI: 0.882-0.887, ASAT (R^2^ = 0.934; 0.932-0.935), and GFAT (R^2^ = 0.932; 0.930-0.934) volumes (**Figure 1B; Supplementary Table S1; Supplementary Figures S2-S5**).^14^ Performance was consistent when the cohort was age-stratified, but attenuated in sex and BMI subgroups, consistent with the marked sex dimorphism of these traits and a high correlation with BMI (**Supplementary Tables S1-S3; Supplementary Figures S6-S7**). Performance was broadly consistent across Black, East Asian, and South Asian participants – with the exception of attenuated VAT prediction in Black participants (R^2^ = 0.784; 95%CI: 0.735-0.823) – although these comparisons were limited by sample size (**Supplementary Table S4**).

### Silhouette-based predictions outperform anthropometric models

We next set out to compare the performance of silhouette-derived predictions of VAT, ASAT, and GFAT volumes with models based on anthropometric measurements. We constructed sex-stratified models combining age with one of – or a combination of – weight, height, body mass index (BMI), waist circumference, hip circumference, waist-to-hip ratio (WHR), and five bioelectric impedance measurements (**Supplementary Table S5**).

BMI-based models offered considerable predictive performance for each fat depot volume, with the poorest performance observed in male participants for prediction of VAT (R^2^ = 0.608; 95% CI: 0.599-0.618) and the best performance observed in female participants for prediction of ASAT (R^2^ = 0.833; 95% CI: 0.828-0.837) (**Figure 2**; **Supplementary Table S6**). These models reflect the high correlation between BMI and any given fat volume in the body (**Supplementary Figure S7**). Silhouette-based models outperformed BMI-based models by ΔR^2^ = 0.220-0.241 for VAT, ΔR^2^ = 0.114-0.172 for ASAT, and ΔR^2^ = 0.248-0.263 for GFAT, suggesting that significant BMI-independent variation in these three fat depots was captured. In contrast, waist circumference-based models displayed only a small improvement for prediction of VAT (R^2^ Male: 0.637; 95% CI: 0.628-0.645; R^2^ Female: 0.659; 95% CI: 0.650-0.667) over BMI-based models and performed worse for the prediction of ASAT and GFAT. Finally, we combined all anthropometric and bioimpedance measures in a single model. While improved performance was observed for VAT (R^2^ Male: 0.724; 95% CI: 0.717-0.732; R^2^ Female: 0.731; 95% CI: 0.723-0.739), ASAT (R^2^ Male: 0.829; 95% CI: 0.823-0.835; R^2^ Female: 0.898; 95% CI: 0.895-0.901), and GFAT (R^2^ Male: 0.793; 95% CI: 0.785-0.801; R^2^ Female: 0.856; 95% CI: 0.852-0.860), silhouette-based models outperformed these models by ΔR^2^ = 0.101-0.125 for VAT, ΔR^2^ = 0.049-0.065 for ASAT, and ΔR^2^ = 0.092-0.098 for GFAT.

**Figure 2.**
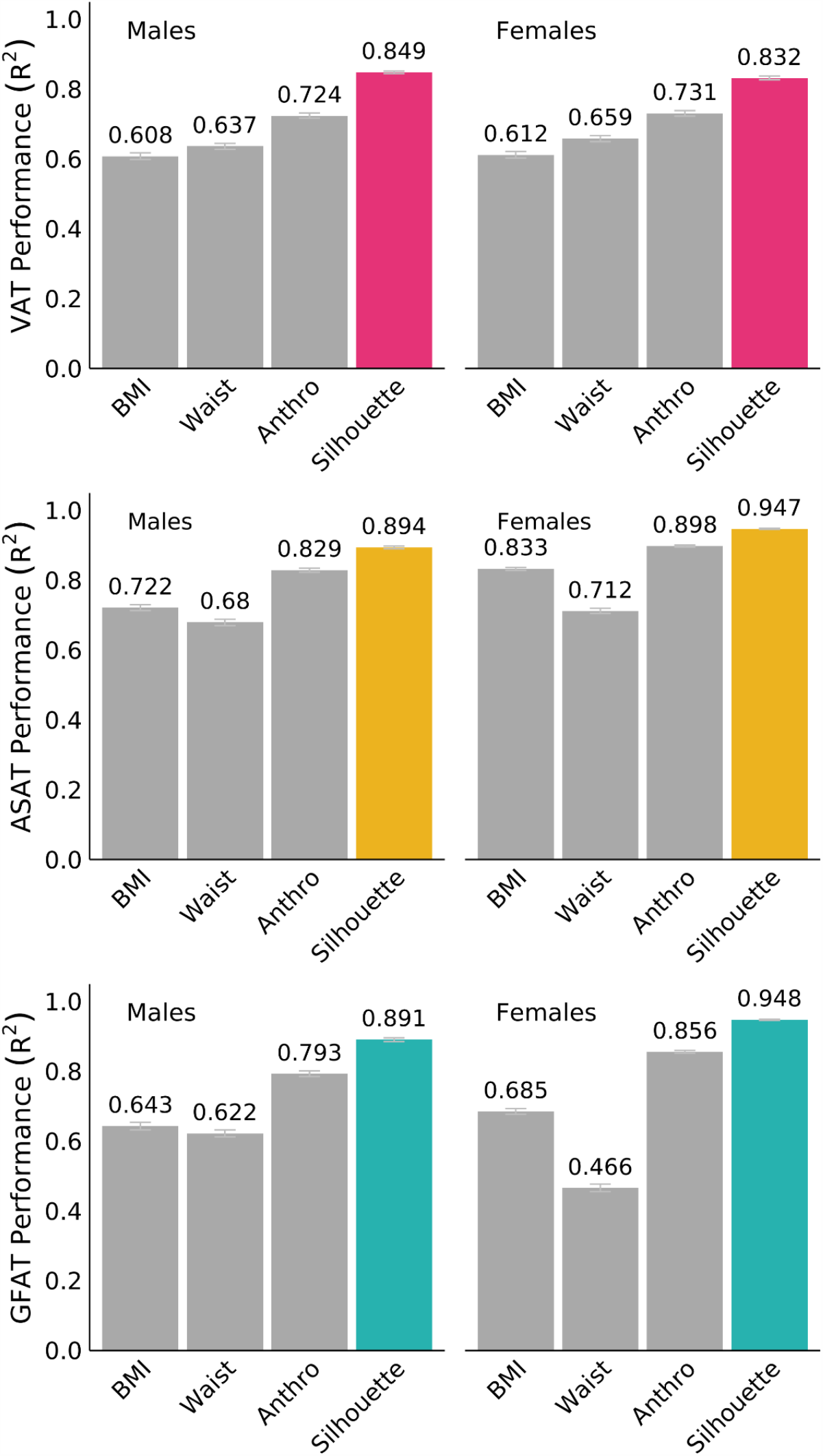
Silhouettes outperform anthropometric models in predicting VAT, ASAT, and GFAT volumes. Sex-specific linear models combining age and various anthropometric metrics measured at the time of imaging were compared to a linear model combining age and silhouette-predicted fat volume (**Supplementary Table S5-S6**). Model definitions; BMI: age + BMI; Waist: age + waist circumference; Anthro: age + weight + height + BMI + waist circumference + hip circumference + waist-hip ratio + body impedance measures; Silhouette: age + silhouette.

We next compared our silhouette-based model for VAT prediction with a recently developed multivariable model for predicting DEXA-derived VAT mass based on 17 anthropometric variables (**Supplementary Table S7**).^34^ These models performed similarly to the combined anthropometric model in this study for VAT prediction (R^2^ Male: 0.719; 95% CI: 0.709-0.728; R^2^ Female: 0.710; 95% CI: 0.694-0.724) – silhouette-based models outperformed these by ΔR^2^ = 0.122-0.130. Taken together, these data suggest that a deep learning model trained on silhouettes can outperform models combining anthropometric and bioimpedance measurements for prediction of VAT, ASAT, and GFAT volumes.

### Silhouette prediction of VAT/ASAT ratio overcomes a key limitation of measured waist circumference

Waist circumference is often used as a proxy for VAT, but the parameter it aims to estimate – central adiposity – can be driven by a preponderence of either ASAT or VAT.^15^ As an example, a pair of age, sex, BMI, and waist circumference-matched participants are shown in **Figure 3A** with highly discordant abdominal fat distribution – one participant has significantly greater VAT (VAT: 9.2 L, ASAT: 4.5 L, VAT/ASAT ratio = 2.0), while the other has much more ASAT (VAT: 3.7 L, ASAT: 9.3 L, VAT/ASAT ratio = 0.40).

**Figure 3.**
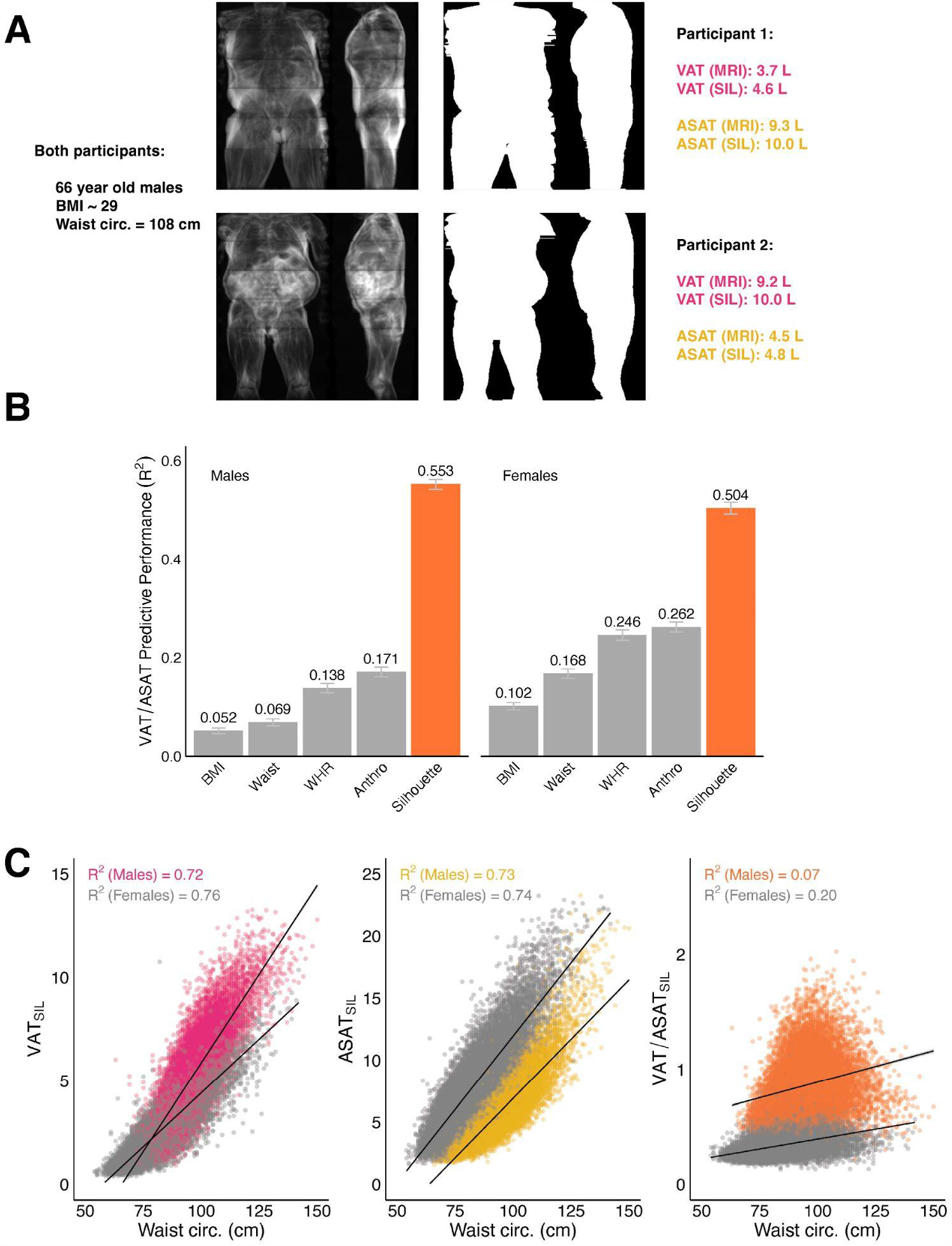
Silhouettes estimate VAT/ASAT ratio, a metric of unfavorable fat distribution. **A)** 2D MRI projections and silhouettes of an age, sex, BMI, and waist circumference matched pair of participants with drastic differences in abdominal fat distribution. While both participants have an elevated waist circumference for their sex- and BMI-group, participant 1 primarily has ASAT-driven central obesity, while participant 2 primarily has VAT-driven central obesity. **B)** A linear model combining age + silhouette prediction markedly outperforms anthropometric models for the prediction of VAT/ASAT ratio (**Supplementary Table S6**). **C)** Waist circumference is strongly correlated with silhouette-predicted VAT (VAT_SIL_) and silhouette-predicted ASAT (ASAT_SIL_) (R^2^ 0.72-0.76), but nearly independent of silhouette-predicted VAT/ASAT (VAT/ASAT_SIL_) (R^2^ 0.07-0.20).

We aimed to investigate the extent to which VAT/ASAT ratio – a marker of unhealthy fat distribution – could be predicted using anthropometric models. In contrast to their performance for fat depot volumes, sex-specific models combining weight, height, BMI, waist circumference, hip circumference, WHR, and five bioimpendance measures yielded poor predictive performance for VAT/ASAT ratio (R^2^ Male: 0.171; 95% CI: 0.161-0.181; R^2^ Female: 0.262; 95% CI: 0.252-0.272) (**Figure 3B, Supplementary Table S6**). Notably, WHR-based models (without other anthropometric measures) achieved comparable performance (R^2^ Male: 0.138; 95% CI: 0.129-0.148; R^2^ Female: 0.246; 95% CI: 0.235-0.256).

This marked reduction in performance compared to similar anthropometric models used to predict VAT, ASAT, and GFAT volumes demonstrates the challenge of predicting regional adiposity out of proportion to the overall size of an individual. Much of the predictive performance for fat depot volumes with variables such as BMI and waist circumference comes from an underlying correlation of all of these variables with the overall size of an individual – VAT/ASAT ratio subverts this pattern, being relatively independent of BMI (r Male = 0.14; r Female = 0.22) (**Supplementary Figure S7**).

We hypothesized that a deep learning model trained on silhouettes could predict VAT/ASAT ratio better than what might be achieved with anthropometric measures, despite the fact that the anatomical boundary between VAT and ASAT cannot be directly ascertained from an individual’s silhouette.

Silhouette-based models demonstrated marked improvement over anthropometric models for prediction of VAT/ASAT ratio (R^2^ Male: 0.553; 95% CI: 0.542-0.562; R^2^ Female: 0.504; 95% CI: 0.492-0.516) (**Figure 3B**). Compared to the best anthropometric model, this represented an improvement of ΔR^2^ = 0.382 in male participants and ΔR^2^ = 0.242 in female participants.

We additionally confirmed that waist circumference was strongly correlated with silhouette-predicted VAT (R^2^ Male 0.72; R^2^ Female 0.76) and silhouette-predicted ASAT (R^2^ Male 0.73; R^2^ Female 0.74), but a poor proxy for silhouette-predicted VAT/ASAT (R^2^ Male 0.07, R^2^ Female 0.20), suggesting that information independent of waist circumference was learned (**Figure 3C, Supplementary Figure S7**).

### Silhouette-predicted VAT/ASAT associates with cardiometabolic diseases

We next investigated associations of silhouette-predicted VAT/ASAT with type 2 diabetes and coronary artery disease (**Supplementary Table S8**).^35^ In sex-specific logistic regression models adjusted for age and imaging center, silhouette-predicted VAT/ASAT was associated with increased prevalence of type 2 diabetes in both males (OR/SD 1.78; 95% CI: 1.69-1.88) and females (OR/SD 1.97; 95% CI: 1.85-2.09) (**Figure 4A**; **Supplementary Table S9**). Additionally adjusting for BMI and waist circumference minimally attenuated effect sizes for silhouette-predicted VAT/ASAT ratio (OR/SD Male 1.70; 95% CI: 1.61-1.80; OR/SD Female 1.74; 95% CI: 1.62-1.86), consistent with the relative independence of VAT/ASAT ratio with these two anthropometric measures. Similar trends were observed with coronary artery disease with attenuated effect sizes – in models adjusted for BMI and waist circumference, silhouette-predicted VAT/ASAT associated with increased prevalence in both males (OR/SD 1.22; 95% CI: 1.16-1.29) and females (OR/SD 1.21; 95% CI: 1.10-1.34).

**Figure 4.**
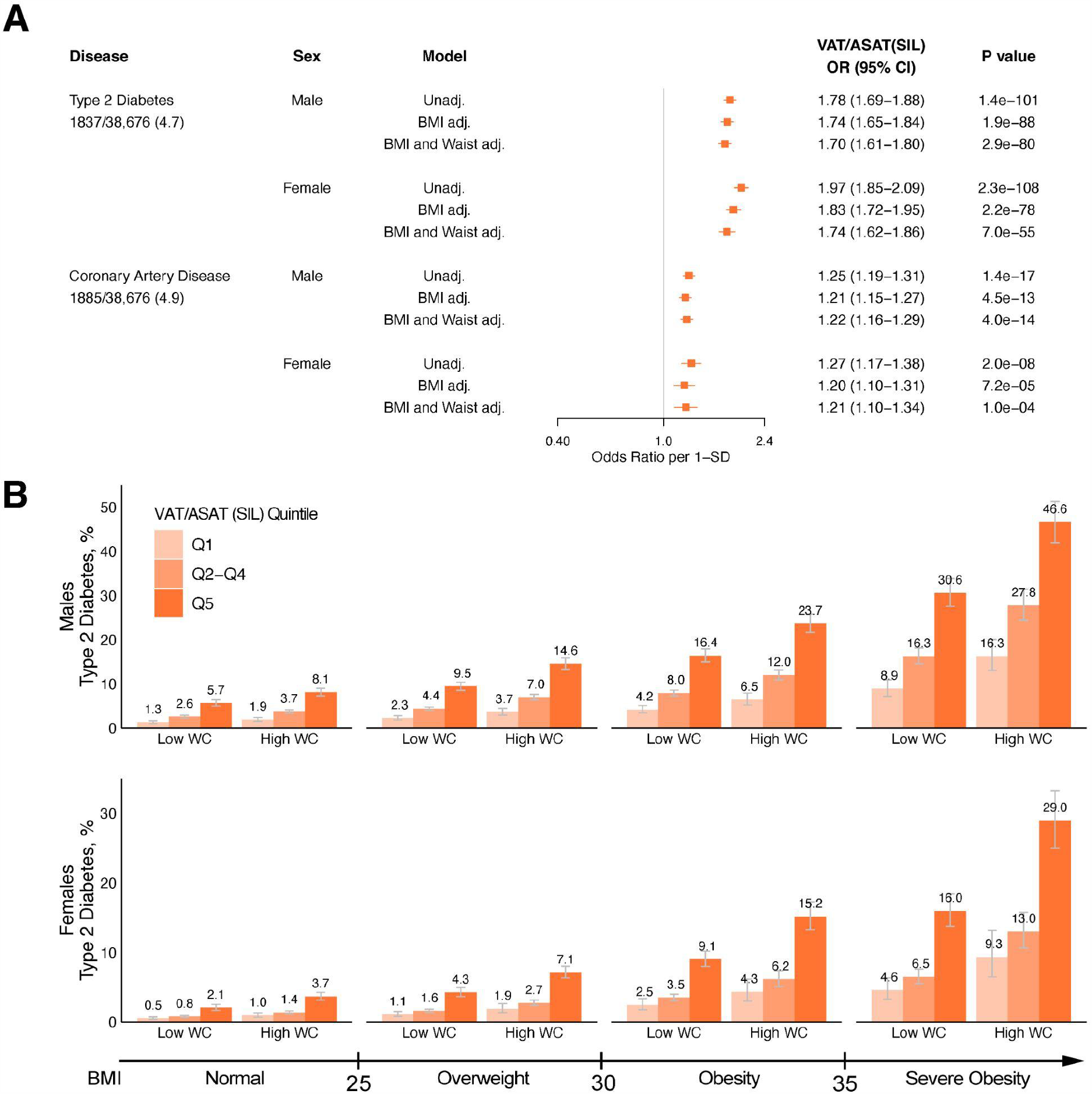
Association of silhouette-predicted VAT/ASAT ratio with type 2 diabetes and coronary artery disease. **A)** Disease associations with silhouette-predicted VAT/ASAT ratio in sex-specific unadjusted and adjusted logistic regression models. All models were adjusted for age at the time of imaging and imaging center - “BMI adj” refers to additional adjustment for BMI, while “BMI and Waist adj.” refers to additional adjustment for BMI and waist circumference. Full data are available in **Supplementary Tables S9 and S12. B)** Sex-stratified standardized prevalence of type 2 diabetes across the bottom quintile (light orange), quintiles 2-4 (neutral orange), and the top quintile (dark orange) of silhouette-predicted VAT/ASAT ratio within BMI-bins and waist circumference categories. High waist circumference was defined in a sex- and BMI-subgroup specific fashion as described in **Supplementary Table S11**.

This procedure was repeated with MRI-derived VAT/ASAT in lieu of silhouette-predicted values to compare disease associations. Trends were broadly consistent between MRI-derived VAT/ASAT and silhouette-predicted VAT/ASAT. Interestingly, the association between MRI-derived VAT/ASAT ratio and type 2 diabetes was slightly attenuated compared to silhouette-predicted values in BMI and waist circumference-adjusted models (OR/SD Male 1.43; 95% CI: 1.36-1.51; OR/SD Female 1.51; 95% CI: 1.40-1.62) (**Supplementary Figure S8**; **Supplementary Table S10**). In contrast, the association with coronary artery disease was nearly identical (OR/SD Male 1.18; 95% CI: 1.12-1.25; OR/SD Female 1.19; 95% CI: 1.08-1.30).

We next set out to understand the gradients of absolute prevalence rates according to quintiles of silhouette-predicted VAT/ASAT. We estimated prevalence rates for males and females separately across clinical BMI categories of normal, overweight, obese, and severely obese participants with either normal or elevated waist circumference based on previously recommended BMI-specific cutoffs (**Supplementary Table S11**).^15,36^ This analysis revealed substantial gradients in prevalence of cardiometabolic diseases according to silhouette-predicted VAT/ASAT quintiles within BMI and waist circumference bins (**Figure 4B; Supplementary Table S12**). For example, men with overweight BMI and normal waist circumference with silhouette-predicted VAT/ASAT in the top quintile had a higher probability of type 2 diabetes (9.5%; 95% CI 8.6-10.4%) compared to both (1) men with overweight BMI and elevated waist circumference with silhouette-predicted VAT/ASAT in the bottom quintile (3.7%; 95% CI 3.0-4.5%) and (2) men with obese BMI and normal waist circumference with silhouette-predicted VAT/ASAT in the bottom quintile (4.2%; 95% CI 3.4-5.1%). Similar trends were observed for coronary artery disease (**Supplementary Figure S9; Supplementary Table S12**).

Over a median follow-up of 1.9 years after imaging, 165 (0.4%) and 393 (1.1%) participants had a new diagnosis of type 2 diabetes or coronary artery disease recorded in the electronic health record. Silhouette-predicted VAT/ASAT associations with incident disease were broadly consistent with prevalent analyses. In BMI- and waist circumference-adjusted Cox regressions, silhouette-predicted VAT/ASAT associated with increased risk of incident type 2 diabetes (HR/SD Male 1.38; 95% CI: 1.14-1.67; HR/SD Female 1.47; 95% CI: 1.24-1.74) and increased risk of incident coronary artery disease in males (HR/SD 1.25; 95% CI: 1.11-1.40) (**Supplementary Table S13**). A null effect was observed for incident coronary artery disease in females, although interpretation was limited by sample size (HR/SD 0.97; 95% CI: 0.79-1.17). Similar effects were observed when MRI-derived VAT/ASAT was used in lieu of silhouette-predicted VAT/ASAT (**Supplementary Table S14**). Taken together, these data support silhouette-predicted VAT/ASAT ratio as a strong, BMI- and waist circumference-independent predictor of cardiometabolic diseases.

## DISCUSSION

In this study, we developed a deep learning model trained on an individual’s silhouette that predicts VAT, ASAT, GFAT, and VAT/ASAT in 40,032 individuals. These silhouette-based predictions are significantly more accurate than those based on anthropometric and bioimpedance measures, particularly for VAT/ASAT ratio, a metric of unhealthy fat distribution.^35^ VAT/ASAT ratio as quantified using silhouette images – largely independent of BMI and waist circumference – was strongly associated with cardiometabolic disease including diabetes and coronary artery disease. These results have at least three implications.

First, deep learning models trained on simple, less data-rich imaging modalities may help close the gap between sophisticated imaging-based markers of adiposity and clinical impact. The present study is proof-of-concept that input data as simple as the outline of an individual is likely to harbor considerably more information about that individual’s fat distribution compared to models that combine several clinical measurements such as BMI and waist circumference. These silhouette-predicted estimates – while crude in comparison to MRI-derived measurements – may be sufficient for cardiometabolic risk screening. Our findings extend prior work focused on prediction of fat-free mass and related measures from DEXA, to measures of fat distribution trained using more sophisticated MRI-based assessment.^19–23^ Recent advances in three-dimensional optical scanners have enabled accurate whole-body surface reconstructions capable of predicting body volumes, waist circumference, and overall fat mass, with some of these products available to the public.^22,24,37–39^ Although accurate and non-invasive, the quantities that are most often predicted by these tools are likely to be highly correlated with BMI, waist circumference, and the overall size of an individual. The present study proposes VAT/ASAT ratio as one useful benchmark for determining how much information has been learned about fat distribution independent of BMI. Alongside advances in smartphone camera technology such as LiDAR (light detection and ranging) and recent evidence that smartphone images can quantify overall body fat, our results lay the scientific foundation for a scalable approach to population health management that incorporates assessment of fat distribution.^28^

Second, waist circumference is more strongly correlated with central obesity than BMI, but is unable to distinguish between VAT and ASAT, indices with differing implications for cardiometabolic risk.^14,15^ Consistent with this limitation, models incorporating anthropometric measurements did not allow for accurate prediction of VAT/ASAT ratio, which we establish as a largely BMI-independent measure of local adiposity. By contrast, our deep learning model showed good predictive performance for VAT/ASAT ratio, even though the boundary between visceral and subcutaneous fat is not explicitly available in silhouette images. The striking gradients in disease patterns observed for VAT/ASAT ratio were minimally attenuated after additional adjustment for BMI and waist circumference. In light of renewed calls for the routine measurement of waist circumference to better stratify cardiometabolic risk associated with body habitus, our work suggests that VAT/ASAT ratio could provide important additional and largely independent information to inform clinical risk estimation.^15^

Third, the conceptual approach outlined here could be leveraged to identify individuals with undiagnosed lipodystrophies or similar phenotypes currently ‘flying under the radar’ within clinical practice.^40^ As an example, familial partial lipodystrophy is a genetic disorder characterized by relative depletion of subcutaneous fat with relative maintenance or excess of visceral fat.^41,42^ Prior proof of principle data suggests that it may be possible to differentiate lipodystrophy patients versus controls using a ‘fat shadow’ derived from clinical-grade DEXA imaging.^42^ Given that this condition remains under recognized within practice, systematic assessment of large populations may prove useful in identifying additional individuals who would benefit from genetic testing or a targeted therapy. For example, metreleptin improves the metabolic profile of patients with partial lipodystrophy and tesamorelin selectively reduces visceral fat in patients with HIV despite no impact on BMI and overall weight.^43–45^ Beyond monogenic lipodystrophies, there is increasing evidence of a less severe, ‘polygenic’ form of lipodystrophy common in individuals with insulin resistance and less pronounced perturbations in fat distribution.^46–48^ Identification of such individuals could, in principle, enable a clinical trial or other assessment of this population to characterize clinical utility.

## STUDY LIMITATIONS

This study has several limitations, providing opportunities for future investigation. First, the majority of participants in the UK Biobank are white, and the imaged substudy investigated here is of mean age 65 years. Although our data suggests similar performance within participant subgroups based on age and ethnicity group, additional validation across ancestrally and geographically diverse populations would be of considerable value, especially given prior evidence of significant variability in fat distribution indices across racial groups.^49^ An important example relates to the South Asian population, where abnormal fat distribution has been postulated as a key driver of the markedly increased rates of cardiovascular disease and diabetes observed, often in the context of a relatively normal BMI.^50,51^ Second, silhouettes in this study were derived by taking the outline of whole-body MRI images, rather than a more cost-effective modality such as photos taken with a smartphone. A future study that utilizes silhouettes obtained from smartphone images would need to additionally account for heterogeneity in user image acquisition technique and require independent validation. Third, we were unable to assess the accuracy of silhouettes in estimating fat depot volume changes over time. Investigation of multiple silhouette-predicted fat depot estimates over time, ideally in the context of a specific lifestyle or clinical intervention, is likely to be of considerable interest.

## CONCLUSIONS

In conclusion, we demonstrate that a deep learning model using silhouettes can quantify fat distribution phenotypes with important potential clinical implications for cardiometabolic health. These results lay the scientific foundation for a population health effort that allows for tracking of these traits in the general population without the requirement for medical imaging.

## Research in context

### Evidence before this study

We searched Pubmed on January 7, 2022 using the terms (“adipose” OR “body composition”) AND (“body shape” OR “silhouette” OR “2D image” OR “3D image” OR “digital image” OR “stereovision” OR “optical imaging”) and no additional restrictions based on date or language. Our search yielded 616 results, of which 32 reported an approach to predict body composition and/or adiposity measures based on a simple input such as a 2D or 3D image. The majority of these studies include up to hundreds of participants, often do not report prediction of specific fat depot volumes, and do not report prediction of fat depot ratios, which are less dependent on the overall size of an individual and so are more challenging to predict. None of the studies reported associations with prevalent or incident cardiometabolic diseases.

### Added value of this study

We set out to understand how well a deep learning model trained on an individual’s silhouette can predict visceral (VAT), abdominal subcutaneous (ASAT), and gluteofemoral (GFAT) adipose tissue volumes and VAT/ASAT ratio in 40,032 individuals. Two-dimensional silhouette images were derived from raw MRI imaging data. Of comparable studies aiming to study the body composition prediction potential of an information-poor imaging input, the present study to the best of our knowledge represents the largest sample size to date, the first to demonstrate prediction of a fat depot ratio (VAT/ASAT), and the first to demonstrate that a model-predicted parameter (in this study, silhouette-predicted VAT/ASAT ratio) can stratify risk of cardiometabolic diseases.

### Implications of all the available evidence

The present study suggests that a data input as simple as the outline of an individual harbors considerable information about the relative fat depot burden in that individual. Predictions from this framework can stratify risk of type 2 diabetes and coronary artery disease independent of body-mass index and waist circumference, suggesting value in obtaining crude estimates of relative fat depot burden.

## Data Availability

The raw UK Biobank data - including the anthropometric data reported here - are made available to researchers from universities and other research institutions with genuine research inquiries following IRB and UK Biobank approval. Representative code used in this work will be deposited at the following Github repository:  https://github.com/broadinstitute/ml4h/tree/master/model_zoo/silhouette_mri.

## Authors’ contributions

Conceptualization: M.D.R.K., S.A., and A.V.K.

Methodology: M.D.R.K. and S.A.

Investigation: M.D.R.K., S.A., and N.D.

Funding acquisition: K.N., P.T.E., A.P., K.N., P.B., A.V.K.

Supervision: P.B. and A.V.K.

Writing: M.D.R.K, S.A., N.D., and A.V.K.

## Ethics committee approval

This analysis of data from the UK Biobank was approved by the Mass General Brigham Institutional Review Board and was performed under UK Biobank application #7089.

## References

1. Kivimäki M, Kuosma E, Ferrie JE, et al. Overweight, obesity, and risk of cardiometabolic multimorbidity: pooled analysis of individual-level data for 120 813 adults from 16 cohort studies from the USA and Europe. Lancet Public Health 2017;2(6):e277–85.

2. Calle EE. Overweight, Obesity, and Mortality from Cancer in a Prospectively Studied Cohort of U.S. Adults. N Engl J Med 2003;14.

3. Anderson MR, Geleris J, Anderson DR, et al. Body Mass Index and Risk for Intubation or Death in SARS-CoV-2 Infection : A Retrospective Cohort Study. Ann Intern Med 2020;173(10):782–90.

4. González-Muniesa P, Mártinez-González M-A, Hu FB, et al. Obesity. Nat Rev Dis Primer 2017;3(1):1–18.

5. Karelis AD, St-Pierre DH, Conus F, Rabasa-Lhoret R, Poehlman ET. Metabolic and Body Composition Factors in Subgroups of Obesity: What Do We Know? J Clin Endocrinol Metab 2004;89(6):2569–75.

6. McLaughlin T, Abbasi F, Lamendola C, Reaven G. Heterogeneity in the prevalence of risk factors for cardiovascular disease and type 2 diabetes mellitus in obese individuals: effect of differences in insulin sensitivity. Arch Intern Med 2007;167(7):642–8.

7. Wildman RP, Muntner P, Reynolds K, et al. The obese without cardiometabolic risk factor clustering and the normal weight with cardiometabolic risk factor clustering: prevalence and correlates of 2 phenotypes among the US population (NHANES 1999-2004). Arch Intern Med 2008;168(15):1617–24.

8. Mathew H, Farr OM, Mantzoros CS. Metabolic health and weight: Understanding metabolically unhealthy normal weight or metabolically healthy obese patients. Metabolism 2016;65(1):73–80.

9. Stefan N, Schick F, Häring H-U. Causes, Characteristics, and Consequences of Metabolically Unhealthy Normal Weight in Humans. Cell Metab 2017;26(2):292–300.

10. Stefan N. Causes, consequences, and treatment of metabolically unhealthy fat distribution. Lancet Diabetes Endocrinol 2020;8(7):616–27.

11. Ashwell M, Cole TJ, Dixon AK. Obesity: new insight into the anthropometric classification of fat distribution shown by computed tomography. Br Med J Clin Res Ed 1985;290(6483):1692–4.

12. Tchernof A, Després J-P. Pathophysiology of Human Visceral Obesity: An Update. Physiol Rev 2013;93(1):359–404.

13. Neeland IJ, Ross R, Després J-P, et al. Visceral and ectopic fat, atherosclerosis, and cardiometabolic disease: a position statement. Lancet Diabetes Endocrinol 2019;7(9):715–25.

14. Agrawal S, Klarqvist MDR, Diamant N, et al. Association of machine learning-derived measures of body fat distribution in &gt;40,000 individuals with cardiometabolic diseases. medRxiv 2021;2021.05.07.21256854.

15. Ross R, Neeland IJ, Yamashita S, et al. Waist circumference as a vital sign in clinical practice: a Consensus Statement from the IAS and ICCR Working Group on Visceral Obesity. Nat Rev Endocrinol 2020;16(3):177–89.

16. Song X, Jousilahti P, Stehouwer CDA, et al. Comparison of various surrogate obesity indicators as predictors of cardiovascular mortality in four European populations. Eur J Clin Nutr 2013;67(12):1298–302.

17. Pischon T, Boeing H, Hoffmann K, et al. General and abdominal adiposity and risk of death in Europe. N Engl J Med 2008;359(20):2105–20.

18. Jacobs EJ, Newton CC, Wang Y, et al. Waist circumference and all-cause mortality in a large US cohort. Arch Intern Med 2010;170(15):1293–301.

19. Xie B, Avila JI, Ng BK, et al. Accurate body composition measures from whole-body silhouettes. Med Phys 2015;42(8):4668–77.

20. Tian IY, Ng BK, Wong MC, et al. Predicting 3D body shape and body composition from conventional 2D photography. Med Phys 2020;47(12):6232–45.

21. Affuso O, Pradhan L, Zhang C, et al. A method for measuring human body composition using digital images. PloS One 2018;13(11):e0206430.

22. Kennedy S, Hwaung P, Kelly N, et al. Optical imaging technology for body size and shape analysis: evaluation of a system designed for personal use. Eur J Clin Nutr 2020;74(6):920–9.

23. Ng BK, Hinton BJ, Fan B, Kanaya AM, Shepherd JA. Clinical anthropometrics and body composition from 3D whole-body surface scans. Eur J Clin Nutr 2016;70(11):1265–70.

24. Ng BK, Sommer MJ, Wong MC, et al. Detailed 3-dimensional body shape features predict body composition, blood metabolites, and functional strength: the Shape Up! studies. Am J Clin Nutr 2019;110(6):1316–26.

25. Sun J, Xu B, Lee J, Freeland-Graves JH. Novel Body Shape Descriptors for Abdominal Adiposity Prediction Using Magnetic Resonance Images and Stereovision Body Images. Obesity 2017;25(10):1795–801.

26. Lee JJ, Freeland-Graves JH, Pepper MR, Yu W, Xu B. Efficacy of thigh volume ratios assessed via stereovision body imaging as a predictor of visceral adipose tissue measured by magnetic resonance imaging. Am J Hum Biol Off J Hum Biol Counc 2015;27(4):445–57.

27. Wang Q, Lu Y, Zhang X, Hahn JK. A Novel Hybrid Model for Visceral Adipose Tissue Prediction using Shape Descriptors. Annu Int Conf IEEE Eng Med Biol Soc IEEE Eng Med Biol Soc Annu Int Conf 2019;2019:1729–32.

28. Majmudar MD, Chandra S, Kennedy S, et al. Smartphone Camera Based Assessment of Adiposity: A Multi-Site Validation Study [Internet]. Public and Global Health; 2021 [cited 2021 Aug 23]. Available from: http://medrxiv.org/lookup/doi/10.1101/2021.06.10.21258595

29. Littlejohns TJ, Holliday J, Gibson LM, et al. The UK Biobank imaging enhancement of 100,000 participants: rationale, data collection, management and future directions. Nat Commun 2020;11(1):2624.

30. Huang G, Liu Z, van der Maaten L, Weinberger KQ. Densely Connected Convolutional Networks. ArXiv160806993 Cs [Internet] 2018 [cited 2021 Apr 20];Available from: http://arxiv.org/abs/1608.06993

31. Sudlow C, Gallacher J, Allen N, et al. UK biobank: an open access resource for identifying the causes of a wide range of complex diseases of middle and old age. PLoS Med 2015;12(3):e1001779.

32. Linge J, Borga M, West J, et al. Body Composition Profiling in the UK Biobank Imaging Study. Obes Silver Spring Md 2018;26(11):1785–95.

33. West J, Leinhard OD, Romu T, et al. Feasibility of MR-Based Body Composition Analysis in Large Scale Population Studies. PLOS ONE 2016;11(9):e0163332.

34. Karlsson T, Rask-Andersen M, Pan G, et al. Contribution of genetics to visceral adiposity and its relation to cardiovascular and metabolic disease. Nat Med 2019;25(9):1390–5.

35. Kaess BM, Pedley A, Massaro JM, Murabito J, Hoffmann U, Fox CS. The ratio of visceral to subcutaneous fat, a metric of body fat distribution, is a unique correlate of cardiometabolic risk. Diabetologia 2012;55(10):2622–30.

36. Ardern CI, Janssen I, Ross R, Katzmarzyk PT. Development of health-related waist circumference thresholds within BMI categories. Obes Res 2004;12(7):1094–103.

37. Heymsfield SB, Bourgeois B, Ng BK, Sommer MJ, Li X, Shepherd JA. Digital anthropometry: a critical review. Eur J Clin Nutr 2018;72(5):680–7.

38. Tinsley GM, Moore ML, Dellinger JR, Adamson BT, Benavides ML. Digital anthropometry via three-dimensional optical scanning: evaluation of four commercially available systems. Eur J Clin Nutr 2020;74(7):1054–64.

39. Tinsley GM, Moore ML, Benavides ML, Dellinger JR, Adamson BT. 3-Dimensional optical scanning for body composition assessment: A 4-component model comparison of four commercially available scanners. Clin Nutr 2020;39(10):3160–7.

40. Gonzaga-Jauregui C, Ge W, Staples J, et al. Clinical and Molecular Prevalence of Lipodystrophy in an Unascertained Large Clinical Care Cohort. Diabetes 2020;69(2):249–58.

41. Shackleton S, Lloyd DJ, Jackson SN, et al. LMNA, encoding lamin A/C, is mutated in partial lipodystrophy. Nat Genet 2000;24(2):153–6.

42. Meral R, Ryan BJ, Malandrino N, et al. “Fat Shadows” From DXA for the Qualitative Assessment of Lipodystrophy: When a Picture Is Worth a Thousand Numbers. Diabetes Care 2018;41(10):2255–8.

43. Oral EA, Gorden P, Cochran E, et al. Long-term effectiveness and safety of metreleptin in the treatment of patients with partial lipodystrophy. Endocrine 2019;64(3):500–11.

44. Sekizkardes H, Cochran E, Malandrino N, Garg A, Brown RJ. Efficacy of Metreleptin Treatment in Familial Partial Lipodystrophy Due to PPARG vs LMNA Pathogenic Variants. J Clin Endocrinol Metab 2019;104(8):3068–76.

45. Stanley TL, Feldpausch MN, Oh J, et al. Effect of Tesamorelin on Visceral Fat and Liver Fat in HIV-Infected Patients With Abdominal Fat Accumulation: A Randomized Clinical Trial. JAMA 2014;312(4):380.

46. Lotta LA, Gulati P, Day FR, et al. Integrative genomic analysis implicates limited peripheral adipose storage capacity in the pathogenesis of human insulin resistance. Nat Genet 2017;49(1):17–26.

47. Lim K, Haider A, Adams C, Sleigh A, Savage DB. Lipodistrophy: a paradigm for understanding the consequences of “overloading” adipose tissue. Physiol Rev 2021;101(3):907–93.

48. Agrawal S, Wang M, Klarqvist MDR, et al. Inherited basis of visceral, abdominal subcutaneous and gluteofemoral fat depots. :46.

49. Kanaley JA, Giannopoulou I, Tillapaugh-Fay G, Nappi JS, Ploutz-Snyder LL. Racial differences in subcutaneous and visceral fat distribution in postmenopausal black and white women. Metabolism 2003;52(2):186–91.

50. Raji A, Seely EW, Arky RA, Simonson DC. Body fat distribution and insulin resistance in healthy Asian Indians and Caucasians. J Clin Endocrinol Metab 2001;86(11):5366–71.

51. Patel AP, Wang M, Kartoun U, Ng K, Khera AV. Quantifying and Understanding the Higher Risk of Atherosclerotic Cardiovascular Disease Among South Asian Individuals: Results From the UK Biobank Prospective Cohort Study. Circulation 2021;144(6):410–22.

