## Supplementary Appendix for "Estimating body fat distribution – a driver of cardiometabolic health – from silhouette images"

Klarqvist*, Agrawal*, et al.

### Supplementary Methods

Preparing silhouettes from whole-body magnetic resonance images

Deep learning to predict fat depot volumes using silhouettes

Description of VAT, ASAT, and GFAT labels used to train deep learning models in this study

**Figure S1** Overview of k-fold cross-validation procedure

**Figure S2-S5** Correlation and Bland-Altman plots for silhouette-estimated VAT, ASAT, GFAT, and VAT/ASAT

**Figure S6** Sex-stratified density plots for fat traits

**Figure S7** Sex-stratified correlogram between anthropometric traits and silhouette-predicted fat depots

**Figure S8** Disease associations with MRI-derived VAT/ASAT ratio in unadjusted and adjusted logistic regression models

**Figure S9** Standardized prevalence of coronary artery disease across quintiles of silhouette-predicted VAT/ASAT

### Supplementary References

### Supplemental Methods

#### Preparing silhouettes from whole-body magnetic resonance images

Whole-body MRI data was preprocessed as previously described.[1] In short, whole-body MRIs were acquired in 6 separate stages with varying resolutions which require preprocessing before merging into 3D volumes. Resolutions ranged from 2.232 x 2.232 x 4.5 $mm^3$ (stages 2-4) to 2.232 x 2.232 x 3.0 $mm^3$ (stage 1). We resampled each series to the highest available resolution (voxel = 2.232 x 2.232 x 3.0 $mm^3$) followed by deduplicating overlapping acquisitions and merging into 3D volumes. There are four available phase acquisitions, (1) in-phase, (2) out-of-phase, (3) water phase, and (4) fat phase. For this study of fat distribution, we used only the fat-phase acquisition to segment axial slices.

Silhouettes were then computed from the resampled axial slices in the merged 3D volume. First, the contrast of axial images was enhanced with contrast limited adaptive histogram equalization (CLAHE) with the parameters limit = 2 and tile size = (8, 8).[2] Next, contrast-enhanced images were thresholded with Otsu's method and all connected components were identified and sorted by their area.[3] There are several situations where multiple segmentations should be returned, such as axial images involving the legs. In contrast, there are situations where multiple segmentations are undesirable, such as when the arms are visible in the axial slices or artifacts are inadvertently segmented. In order to distinguish between these two cases, we employed a heuristic such that if the ratio between the top two largest segmentations are >= 0.25 they were both kept and returned. In all other cases, only the largest segmentation was returned. Next, returned segmentations were flood filled such that their interior was completely white. Stacking all these segmentations result in a 3D volume of equal size as the input 3D volume of MRI images.

Coronal and sagittal two-dimensional projections were generated by computing the mean intensity projection in each orientation of the segmented 3D volume. For example, a given pixel on a coronal two-dimensional projection represents the mean intensity across all pixels making up a line oriented in the anterior-posterior direction perpendicular to the coronal plane. This procedure results in a surface map of each participant. Pixel intensities were rounded and then converted into zeros for background and ones for body. The final coronal and sagittal silhouettes were then concatenated side-by-side and resized to 237 x 256 pixels for downstream applications.

#### Deep learning to predict fat depot volumes using silhouettes

DenseNets are constructed with two principal building blocks: (1) dense blocks comprising of batch normalization, the non-linear ReLU activation function, and 3x3 convolutions of increasing number of channels that are propagated from previous layers to enable efficient gradient flow; and (2) transition blocks that compress the number of channels by half using channel-wise convolutions (1x1), and perform a spatial reduction by a factor of 2 by using an average pooling layer of stride 2 and pool size 2. In our model, the channel output of the last dense block convolution was flattened using a global average pooling layer and then fed into a 512-dimensional fully connected layer that then split into three arms, one for each fat depot (VAT, ASAT, and GFAT).[4] Each arm comprised of two fully connected layers of size 128 and 32, with the ReLu non-linearity as their activation functions, followed by a single-dimensional linear output layer. For VAT/ASAT, the 32-dimensional fully connected layers from the VAT and ASAT arms were concatenated resulting in a 64-dimensional latent space that was processed as above with two fully connected layers of size 128 and 32 followed by a linear regression output.

The inputs for this model were the coronal and sagittal binary silhouettes placed side-by-side with the shape 237 x 256 x 1. The models were trained with the Adam optimizer[5] with a learning rate set to a cosine decay policy decaying from 0.0001 to 0 over 100 epochs, weight decay of 0.0001, shrinkage loss[6] with the hyperparameters a = 10.0 and c = 0.2 as the loss function, and a batch size of 32. No additional hyperparameter search or ablation studies were performed.

For all training data, the following augmentations (random permutations of the training images) were applied: random shifts in the XY-plane by up to ±16 pixels and rotations by up to ±5 degrees around its center axis.

**Description of VAT, ASAT, and GFAT labels used to train deep learning models in this study**

A full description of the machine learning methods used to predict VAT, ASAT, and GFAT volumes including performance metrics and associations with type 2 diabetes and coronary artery disease is available in a prior manuscript.[1]

Among UK Biobank participants who underwent MRI imaging study, visceral adipose tissue (VAT) volume, abdominal subcutaneous adipose tissue (ASAT) volume, and total adipose tissue between the bottom of the thigh muscles to the top of vertebrae T9 (TAT) volume were quantified and made available via the UK Biobank portal to the broader research community in a subset of participants.[7–12] VAT (field 22407, "volume of the adipose tissue within the abdominal cavity, excluding adipose tissue outside the abdominal skeletal muscles and adipose tissue and lipids within and posterior of the spine and posterior of the back muscles") was available in 9,978 participants, ASAT (field 22408, "volume of the subcutaneous adipose tissue in the abdomen from the top of the femoral head to the top of the thoracic vertebrae T9") was available in 9,979, and TAT (field 22415, "total volume of adipose tissue, measured by MRI, between the bottom of the thigh muscles to the top of vertebrae T9") was available in 8,524. Based on these definitions, we computed gluteofemoral adipose tissue (GFAT) volume: GFAT = TAT (between top of T9 and bottom of thigh muscles) - VAT - ASAT.

Given that the vast majority of adipose tissue between the top of vertebrae T9 and the top of the femoral head is accounted for by VAT or ASAT, GFAT was defined as total adipose tissue between the top of the femoral head and the bottom of the thigh muscles.

To train convolutional neural network models to measure VAT, ASAT, and GFAT, we first simplified the three-dimensional MRI images into composite two-dimensional projections of coronal and sagittal views, leading to an 830-fold reduction in data input size (**Supplementary Figure SM1**). These machine learning models – trained on 80% of the participants with fat depots previously quantified –

demonstrated near-perfect estimation association of each fat depot in the 20% of remaining individuals for each depot ($r^2$ = 0.991, 0.991, and 0.978 for VAT, ASAT, and GFAT, respectively).

Finally, we applied these models on the remaining unlabeled participants of the UK Biobank imaging substudy (i.e. participants with raw MRI imaging available, but no fat labels returned to the UK Biobank) to compute VAT, ASAT, and GFAT volumes. All participants (including those who had labels returned to the UK Biobank and those who received labels from the machine learning model) were included as truth labels for training of silhouette-based deep learning models in this study.

**Supplementary Figure SM1** Convolutional neural networks to quantify adipose tissue depots from body MRI images

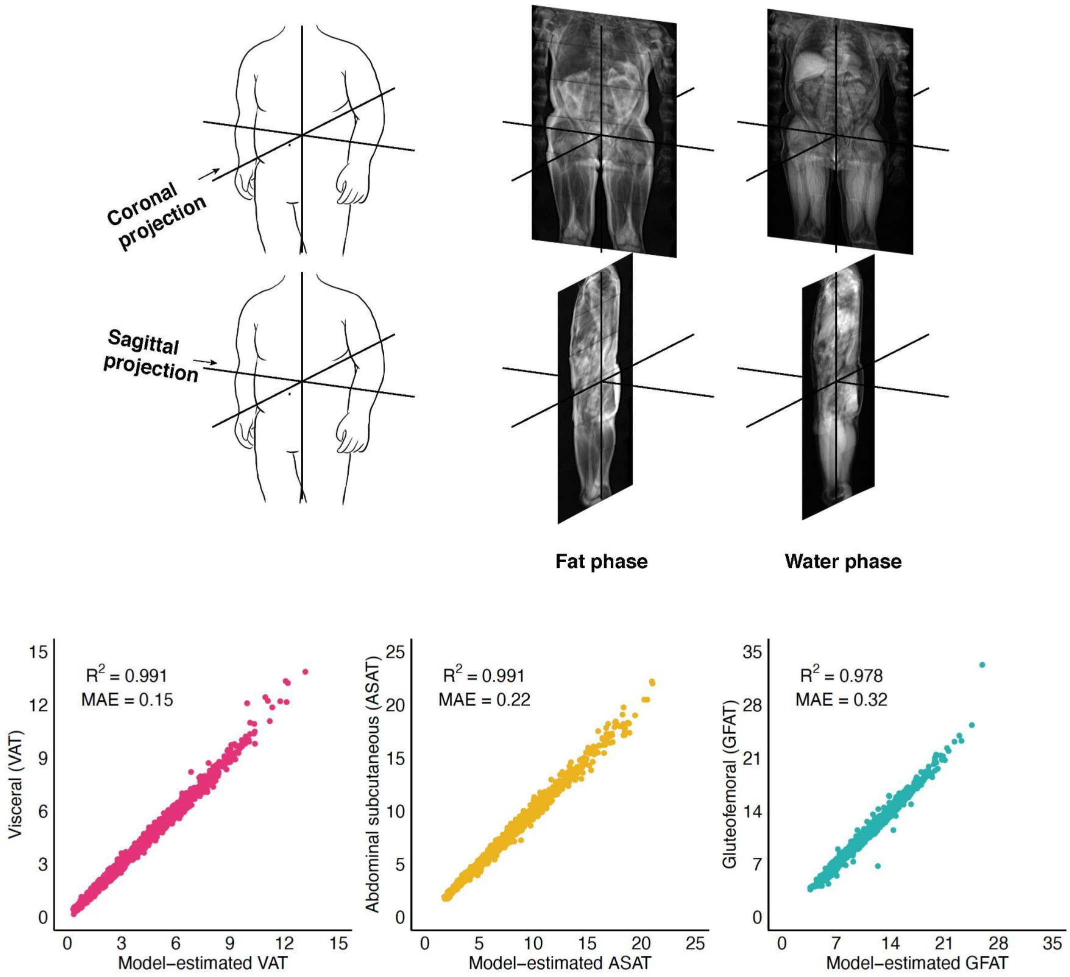


(top row) Sample input into convolutional neural network (CNN): two-dimensional projections of MRIs in the coronal and sagittal directions with fat and water phases are used as input for each individual. (bottom row) In a 20% holdout set among each pre-labeled fat depot, the CNN achieves near-perfect prediction of that fat depot.

**Figure S1 Overview of k-fold cross-validation procedure**

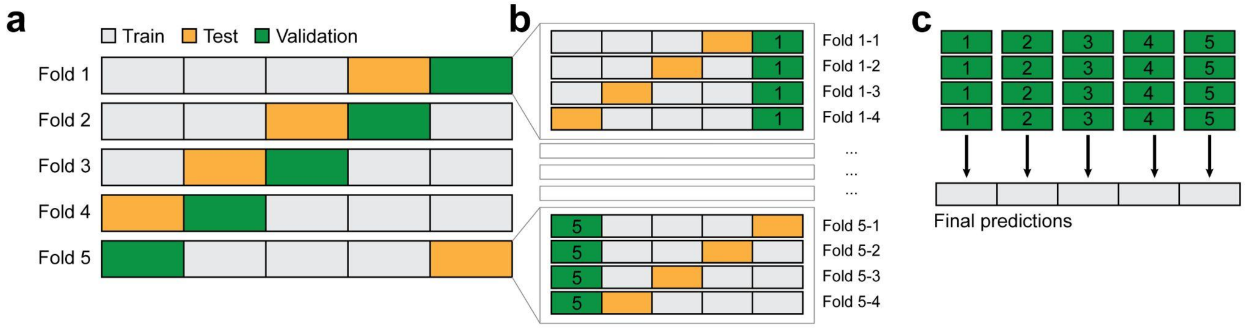


**a)** Five-fold cross-validation with one fold allocated for testing, one for validation, and the remaining three for training. **b)** In a nested cross-validation approach, k-1 models can be trained for each fold when the validation fold is fixed. **c)** For the final predictions, the k-1 models for each k are mean-ensembled.

**Figure S2 Correlation and Bland-Altman plots for silhouette-estimated VAT**

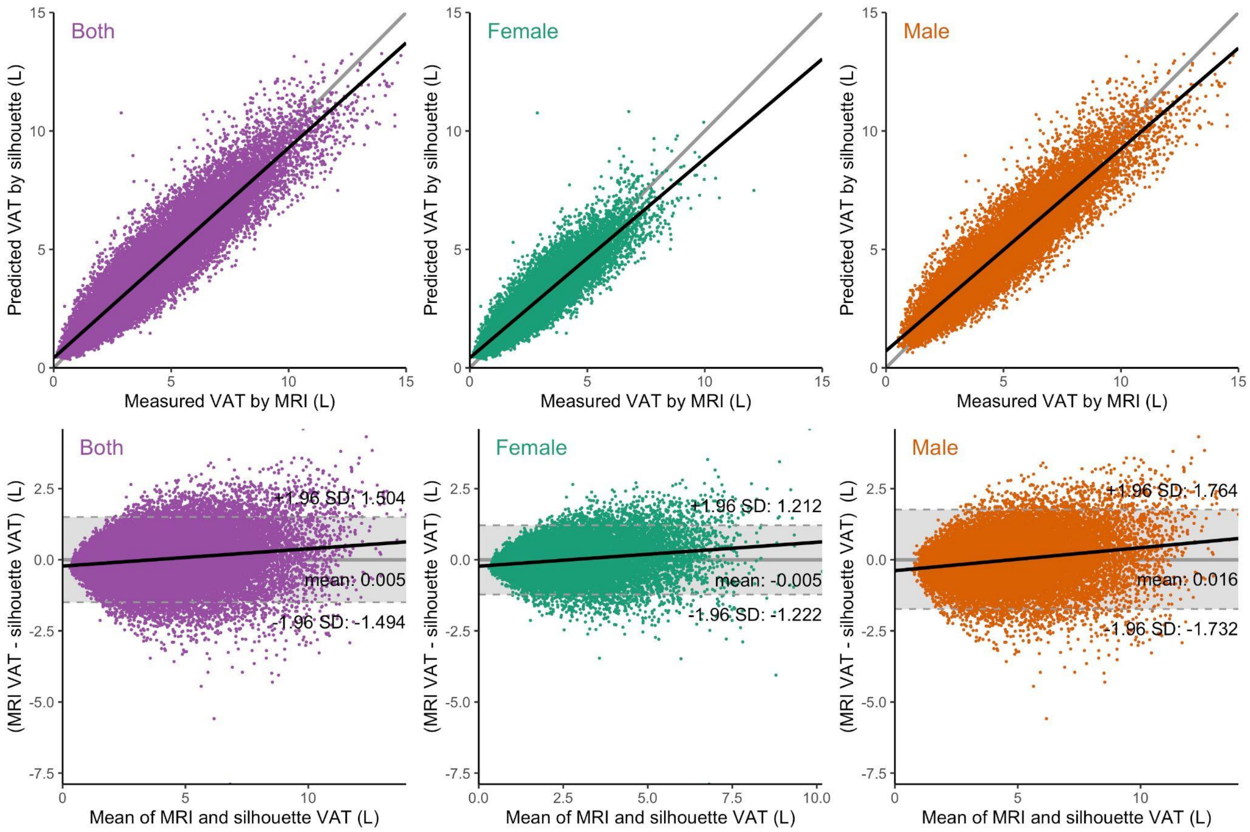


**Upper row:** Silhouette-predicted VAT volume plotted against MRI-measured VAT volume in sex-combined (left), female (middle), and male (right) groups. The black lines denote the linear fits (ordinary least squares regression) and the gray line is the identity function.

**Lower row:** Bland-Altman plots analyze the agreement between two different measurement techniques by showing the mean difference between the approaches on the X-axis and the measurement difference on the Y-axis. The black lines denote the linear fits (ordinary least squares regression) and the three dashed gray lines correspond to ±1.96 standard deviations (SD, 95%) of the difference and the mean difference, respectively. This span between the upper and lower confidence intervals is called the 'limit of agreement' and is shaded in gray. The thicker gray horizontal line shows the ideal mean difference of 0 (no difference).

**Figure S3 Correlation and Bland-Altman plots for silhouette-estimated ASAT**

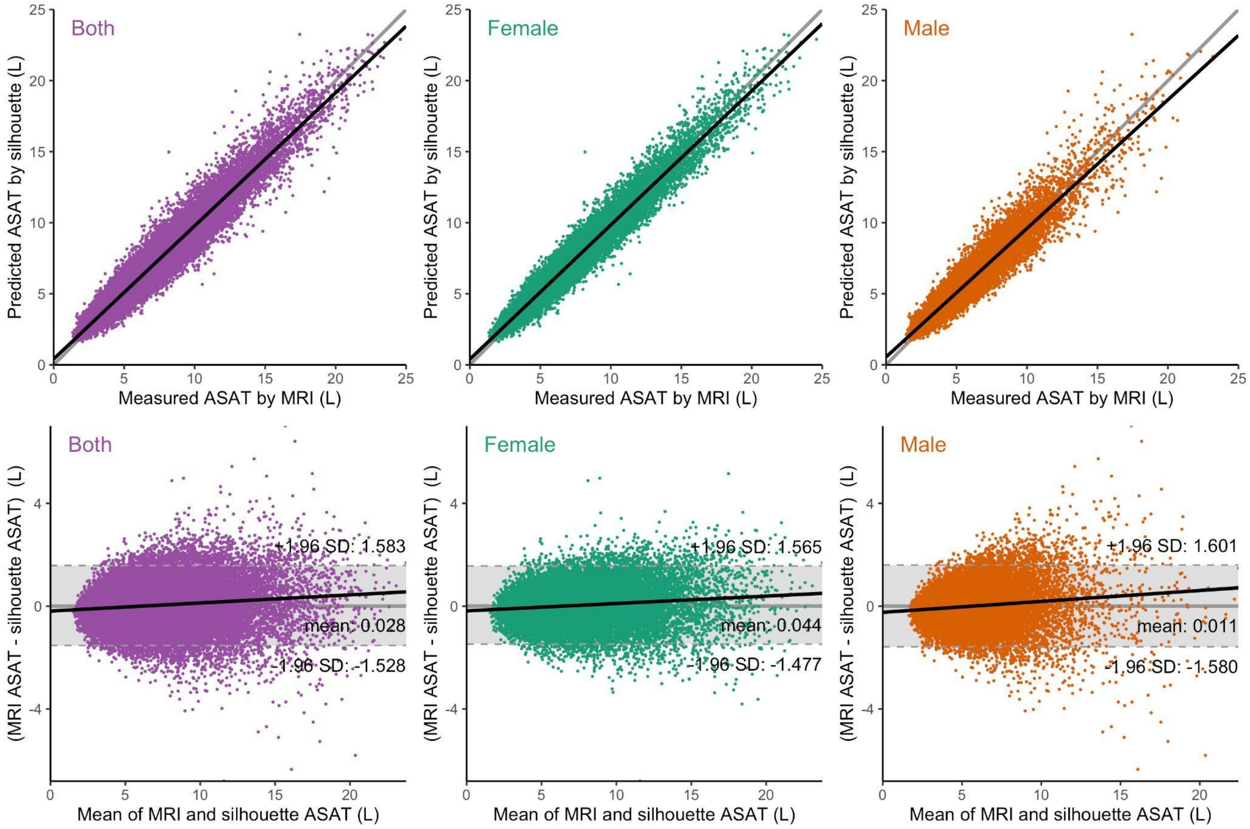


**Upper row:** Silhouette-predicted ASAT volume plotted against MRI-measured ASAT volume in sex-combined (left), female (middle), and male (right) groups. The black lines denote the linear fits (ordinary least squares regression) and the gray line is the identity function.

**Lower row:** Bland-Altman plots analyze the agreement between two different measurement techniques by showing the mean difference between the approaches on the X-axis and the measurement difference on the Y-axis. The black lines denote the linear fits (ordinary least squares regression) and the three dashed gray lines correspond to ±1.96 standard deviations (SD, 95%) of the difference and the mean difference, respectively. This span between the upper and lower confidence intervals is called the 'limit of agreement' and is shaded in gray. The thicker gray horizontal line shows the ideal mean difference of 0 (no difference).

**Figure S4 Correlation and Bland-Altman plots for silhouette-estimated GFAT**

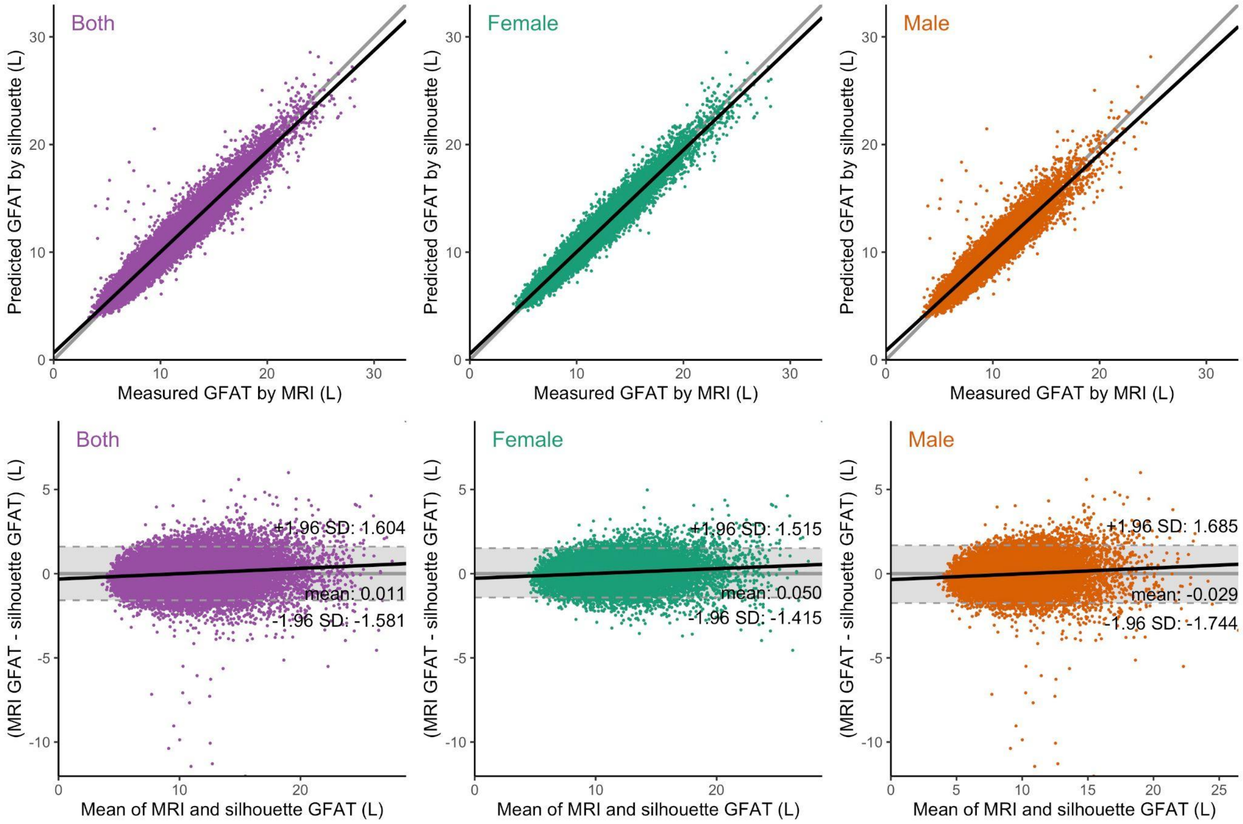


**Upper row:** Silhouette-predicted GFAT volume plotted against MRI-measured GFAT volume in sex-combined (left), female (middle), and male (right) groups. The black lines denote the linear fits (ordinary least squares regression) and the gray line is the identity function.

**Lower row:** Bland-Altman plots analyze the agreement between two different measurement techniques by showing the mean difference between the approaches on the X-axis and the measurement difference on the Y-axis. The black lines denote the linear fits (ordinary least squares regression) and the three dashed gray lines correspond to ±1.96 standard deviations (SD, 95%) of the difference and the mean difference, respectively. This span between the upper and lower confidence intervals is called the 'limit of agreement' and is shaded in gray. The thicker gray horizontal line shows the ideal mean difference of 0 (no difference).

**Figure S5 Correlation and Bland-Altman plots for silhouette-estimated VAT/ASAT**

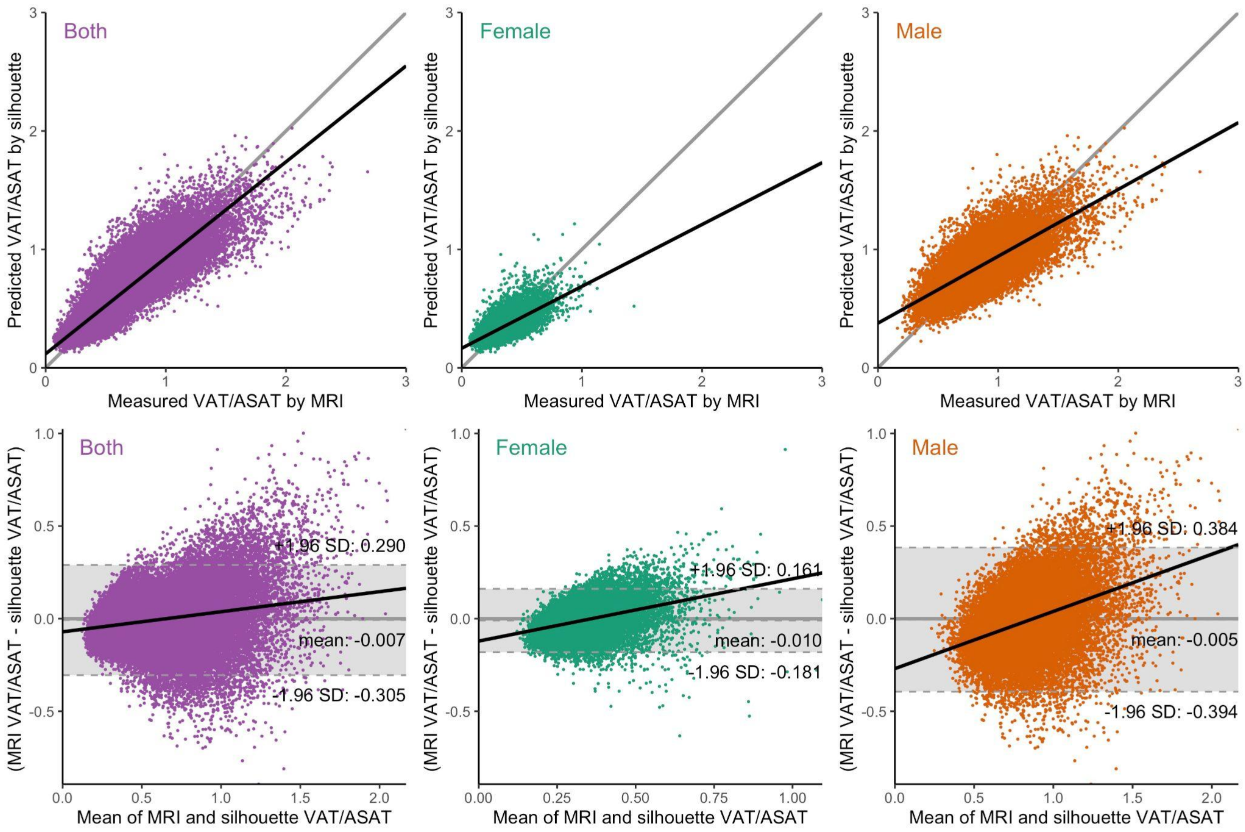


**Upper row:** Silhouette-predicted VAT/ASAT plotted against MRI-measured VAT/ASAT in sex-combined (left), female (middle), and male (right) groups. The black lines denote the linear fits (ordinary least squares regression) and the gray line is the identity function.

**Lower row:** Bland-Altman plots analyze the agreement between two different measurement techniques by showing the mean difference between the approaches on the X-axis and the measurement difference on the Y-axis. The black lines denote the linear fits (ordinary least squares regression) and the three dashed gray lines correspond to ±1.96 standard deviations (SD, 95%) of the difference and the mean difference, respectively. This span between the upper and lower confidence intervals is called the 'limit of agreement' and is shaded in gray. The thicker gray horizontal line shows the ideal mean difference of 0 (no difference).

**Figure S6 Sex-stratified density plots for fat traits**

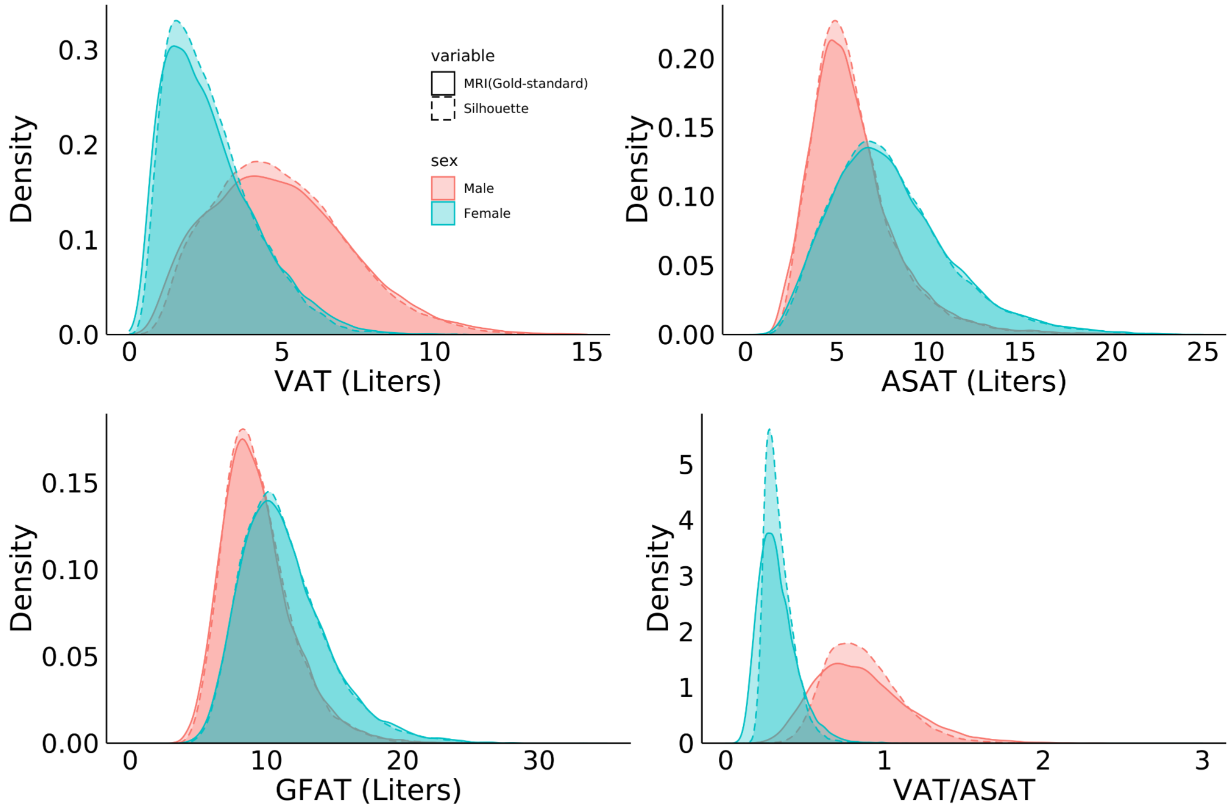


A deep learning model trained on silhouettes is well-calibrated compared to MRI-measured visceral (VAT), abdominal subcutaneous (ASAT), and gluteofemoral (GFAT) adipose tissue volumes, as well as VAT/ASAT ratio.

**Figure S7 Sex-stratified correlogram between anthropometric traits and silhouette-predicted fat depots**

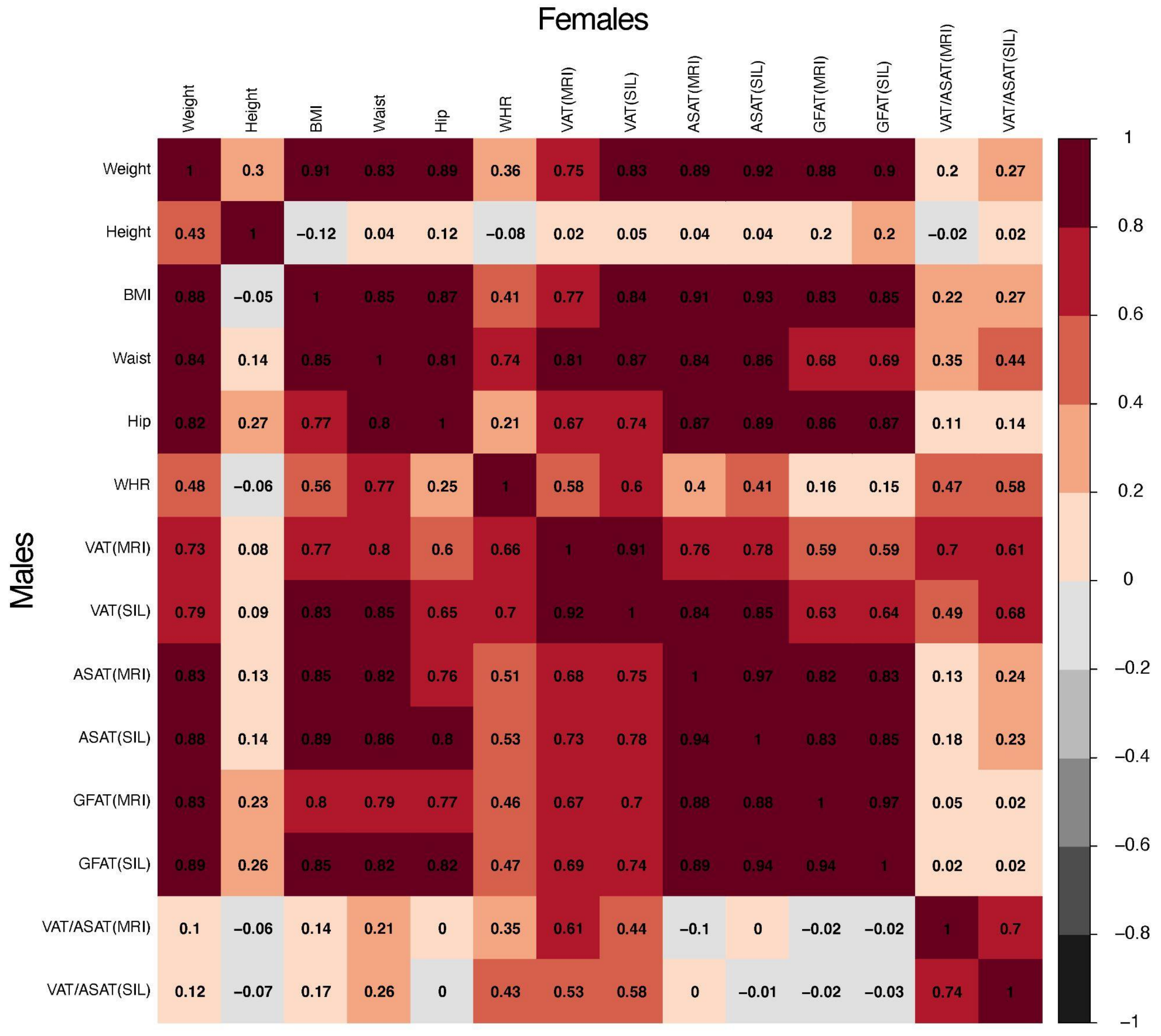


Abbreviations: BMI - body mass index; Waist - waist circumference; Hip - hip circumference; WHR - waist-to-hip ratio; VAT - visceral adipose tissue volume; ASAT - abdominal subcutaneous adipose tissue volume; GFAT - gluteofemoral adipose tissue volume; MRI - gold-standard measurement used as truth label in this study; SIL - output from deep learning model trained on silhouettes discussed in this study.

**Figure S8 Disease associations with MRI-derived VAT/ASAT ratio in unadjusted and adjusted logistic regression models**

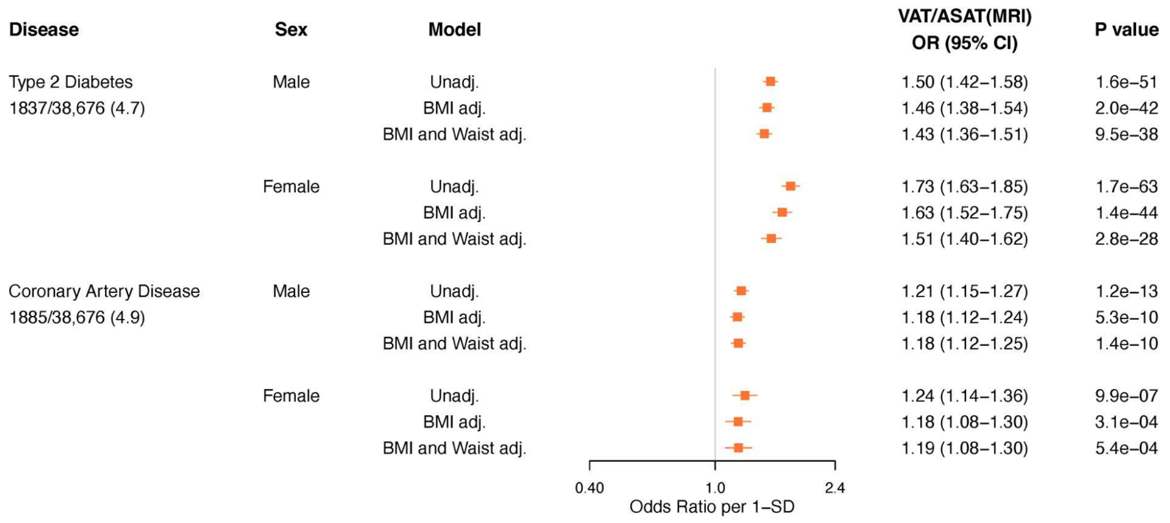


Disease associations with MRI-derived VAT/ASAT ratio in unadjusted, BMI-adjusted, and BMI- and waist circumference-adjusted logistic regression models. All models were adjusted for age at the time of imaging, sex, and imaging center. Full data are available in **Supplementary Table S10**.

**Figure S9 Standardized prevalence of coronary artery disease across quintiles of silhouette-predicted VAT/ASAT**

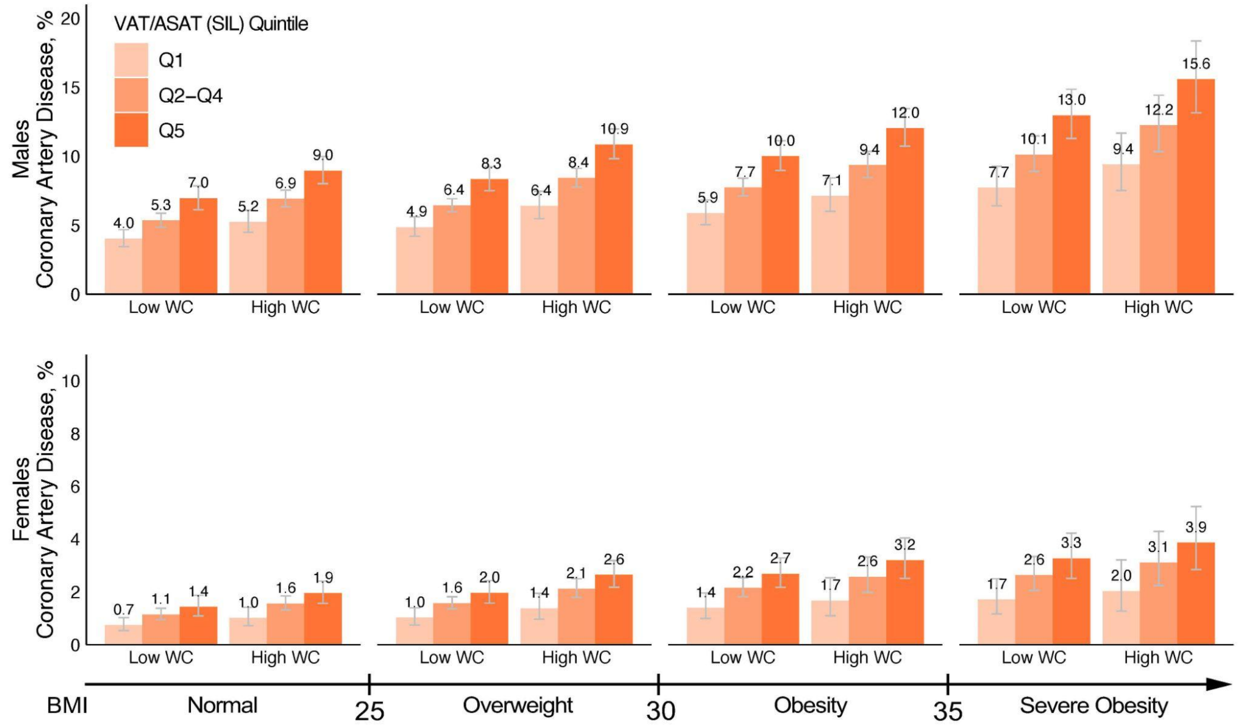


Sex-stratified standardized prevalence of coronary artery disease across the bottom quintile (light orange), quintiles 2-4 (neutral orange), and the top quintile (dark orange) of silhouette-predicted VAT/ASAT ratio within BMI-bins and waist circumference categories. High waist circumference was defined in a sex- and BMI-subgroup specific fashion as described in **Supplementary Table S12**.

**Supplemental References**


1. Agrawal S, Klarqvist MDR, Diamant N, et al. Association of machine learning-derived measures of body fat distribution in >40,000 individuals with cardiometabolic diseases. medRxiv 2021;2021.05.07.21256854.
2. Pizer SM, Amburn EP, Austin JD, et al. Adaptive histogram equalization and its variations. Comput Vis Graph Image Process 1987;39(3):355–68.
3. Otsu N. A Threshold Selection Method from Gray-Level Histograms. IEEE Trans Syst Man Cybern 1979;9(1):62–6.
4. Lin M, Chen Q, Yan S. Network In Network. ArXiv13124400 Cs [Internet] 2014 [cited 2021 Aug 3];Available from: http://arxiv.org/abs/1312.4400
5. Kingma DP, Ba J. Adam: A Method for Stochastic Optimization. ArXiv14126980 Cs [Internet] 2017 [cited 2021 Apr 20];Available from: http://arxiv.org/abs/1412.6980
6. Lu X, Ma C, Ni B, Yang X, Reid I, Yang M-H. Deep Regression Tracking with Shrinkage Loss [Internet]. 2018 [cited 2021 Apr 20]. p. 353–69.Available from: https://openaccess.thecvf.com/content_ECCV_2018/html/Xiankai_Lu_Deep_Regression_Tracking_ECCV_2018_paper.html
7. Leinhard OD, Johansson A, Rydell J, et al. Quantitative abdominal fat estimation using MRI. In: 2008 19th International Conference on Pattern Recognition. 2008. p. 1–4.
8. Borga M, Thomas EL, Romu T, et al. Validation of a fast method for quantification of intra-abdominal and subcutaneous adipose tissue for large-scale human studies. NMR Biomed 2015;28(12):1747–53.
9. West J, Leinhard OD, Romu T, et al. Feasibility of MR-Based Body Composition Analysis in Large Scale Population Studies. PLOS ONE 2016;11(9):e0163332.
10. Borga M, West J, Bell JD, et al. Advanced body composition assessment: from body mass index to body composition profiling. J Investig Med Off Publ Am Fed Clin Res 2018;66(5):1–9.
11. Linge J, Borga M, West J, et al. Body Composition Profiling in the UK Biobank Imaging Study. Obes Silver Spring Md 2018;26(11):1785–95.
12. Linge J, Whitcher B, Borga M, Dahlqvist Leinhard O. Sub-phenotyping Metabolic Disorders Using Body Composition: An Individualized, Nonparametric Approach Utilizing Large Data Sets. Obes Silver Spring Md 2019;27(7):1190–9.
